# Generative Artificial Intelligence Model for Simulating Brain Structural Changes in Schizophrenia

**DOI:** 10.1101/2024.05.29.24308097

**Authors:** Hiroyuki Yamaguchi, Genichi Sugihara, Masaaki Simizu, Yuichi Yamashita

## Abstract

**Background:** Recent advancements in generative artificial intelligence (AI) for image generation have presented significant opportunities for medical imaging, offering a promising avenue for generating realistic virtual medical images while ensuring patient privacy. The generation of a large number of virtual medical images through AI has the potential to augment training datasets for discriminative AI models, particularly in fields with limited data availability, such as neuroimaging. Current studies on generative AI in neuroimaging have mainly focused on disease discrimination; however, its potential for simulating complex phenomena in psychiatric disorders remains unknown. In this study, as examples of a simulation, we aimed to present a novel generative AI model that transforms magnetic resonance imaging (MRI) images of healthy individuals into images that resemble those of patients with schizophrenia (SZ) and explore its application.

**Methods:** We used anonymized public datasets from the Center for Biomedical Research Excellence (SZ, 71 patients; healthy subjects [HSs], 71 patients) and the Autism Brain Imaging Data Exchange (autism spectrum disorder [ASD], 79 subjects; HSs, 105 subjects). We developed a model to transform MRI images of HSs into MRI images of SZ using cycle generative adversarial networks. The efficacy of the transformation was evaluated using voxel-based morphometry to assess the differences in brain region volume and the accuracy of age prediction pre- and post-transformation. In addition, the model was examined for its applicability in simulating disease comorbidities and disease progression.

**Results:** The model successfully transformed HS images into SZ images and identified brain volume changes consistent with existing case-control studies. We also applied this model to ASD MRI images, where simulations comparing SZ with and without ASD backgrounds highlighted the differences in brain structures due to comorbidities. Furthermore, simulation of disease progression while preserving individual characteristics showcased the model’s ability to reflect realistic disease trajectories.

**Discussion:** The findings suggest that our generative AI model can capture subtle changes in brain structures associated with SZ and offers a novel tool for visualizing brain alterations across various conditions. The potential of this model extends beyond clinical diagnoses to advancements in the simulation of disease mechanisms, which may ultimately contribute to the refinement of therapeutic strategies.

## 1 Introduction

Rapid advancements in image generative artificial intelligence (AI) have marked the beginning of new possibilities in various fields (1). Significant breakthroughs include the emergence of DALL-E (2) and stable diffusion (3), which have made the potential of AI for generating realistic and complex images widely known (4). This evolution has profound implications, particularly in the intricate landscape of medical imaging, where concerns regarding privacy, ethics, and legal constraints have historically constrained the sharing of patient data.

The utilization of generative AI models has demonstrated realistic and comprehensive potential for generating two-dimensional medical images, such as chest radiographs and fundus photography (5), and three-dimensional (3D) medical images, including magnetic resonance imaging (MRI) of the brain, chest, and knees (6). These studies have highlighted the potential of generative AI for synthesizing authentic medical images without compromising the confidentiality of sensitive patient information.

The limitation of available medical images, in contrast to the abundance of natural images, emphasizes the importance of generative AI, which facilitates the use of large amounts of labeled data in model training. In neuroimaging, the generative AI approach has been used to generate brain MRI (7), single-photon emission tomography (SPECT) (8), and positron emission tomography (PET) (9). Among these, generative AI is commonly used in medical imaging to improve the performance of models by generating a large number of images and using them as training data, that is, for data augmentation (10–12). It is difficult to increase the number of samples for MRI of actual psychiatric and neurological disorders. Hence, a strategy using generative deep learning techniques, such as generative adversarial networks (GANs), is used to enhance the learning process by expanding the sample size (11,13–15). Zhou et al. demonstrated that a GAN framework could be developed to generate brain MRI images to enhance performance and improve accuracy in classifying Alzheimer’s disease and mild cognitive impairment (16). Zhao et al. introduced a functional network connectivity-based GAN to distinguish patients with schizophrenia (SZ) from healthy subjects (HSs) using functional MRI data (17). Generative AI has also demonstrated strength in neuroimaging (18) segmentation. Furthermore, it is imperative to investigate the efficacy of style transfers derived from generative AI. Style transfer involves applying the characteristics or style of one image to another while preserving the content of the latter. This technique holds promise for transforming easily obtainable images, such as computed tomography scans, into images exclusive to a limited number of facilities, such as MRI scans (19). In addition, the application of the style transfer technique is expected to effectively reduce bias in image quality caused by differences in imaging equipment and sites (20), which are unavoidably included in MRI images (21).

Existing neuroimaging research using generative AI has concentrated primarily on contributions to specific areas, such as disease diagnosis and lesion detection. However, the use of these techniques to simulate more complex clinical phenomena presents an interesting area for further exploration of the potential use of generative AI in neuroimaging research (22). Examples from external medicine include attempts to simulate automated automobile driving (23) and the design of novel proteins (24).

Previous studies provide an example of one such attempt to apply generative AI to psychiatric brain imaging research, as an example of an approach to the problem of comorbidity and heterogeneity in psychiatric disorders (25,26). Specifically, these studies have focused on the relationship between SZ and autism spectrum disorder (ASD). SZ and ASD are defined as distinct disorder groups based on diagnostic criteria but share some common features, such as difficulties in social interaction and communication (27). Furthermore, common features in brain structures and genetic alterations have been noted (28,29). Owing to the complexity of their etiology, the relationship between these two disorders and their comorbid phenotypes remains to be elucidated. The ability of generative AI to simulate these conditions can shed light on the intricate relationship between these disorders and their overlapping phenotypes.

The first step in this study was to develop a generative AI that simulates brain volume changes caused by SZ, specifically a generative AI using CycleGAN. This enables the transformation of brain images from healthy individuals into images similar to those of patients diagnosed with SZ. We validated this artificial schizophrenic brain simulator by analyzing specific brain regions affected by transformation and comparing them with existing findings on SZ. Furthermore, we aimed to evaluate the feasibility of our “SZ brain generator” in simulations of the disease process of SZ and in simulation experiments of the comorbidity of ASD and SZ.

## 2 Materials and Methods

### 2.1 Dataset description

In this study, we used the Center for Biomedical Research Excellence (COBRE; http://fcon_1000.projects.nitrc.org/indi/retro/cobre.html) dataset, which is anonymized and publicly available. All the subjects were diagnosed and screened using the Structured Clinical Interview for the Diagnostic and Statistical Manual of Mental Disorders, 4th edition Axis I Disorders (SCID) (30)(31). Individuals with a history of head trauma, neurological illness, serious medical or surgical illness, or substance abuse were excluded. We selected 142 subjects from this database, including 71 patients with SZ and 71 HSs.

We also used the Autism Brain Imaging Data Exchange (ABIDE; http://fcon_1000.projects.nitrc.org/indi/abide/) dataset, which is a multicenter project that focuses on ASD. It includes > 1000 ASD and typically developing (TD) subjects. The New York City University dataset was used in this study. Finally, we included 184 subjects from this dataset: 79 subjects with ASD and 105 TD subjects.

The demographic and clinical characteristics of the COBRE and ABIDE datasets are presented in Supplementary Table 1.

This study was conducted in accordance with the current Ethical Guidelines for Medical and Health Research Involving Human Subjects in Japan and was approved by the Committee on Medical Ethics of the National Center of Neurology and Psychiatry.

### 2.3 Data preprocessing

MRI data were preprocessed using the Statistical Parametric Mapping software (SPM12, Wellcome Department of Cognitive Neurology, London, UK, https://www.fil.ion.ucl.ac.uk/spm/software/spm12/) with the Diffeomorphic Anatomical Registration Exponentiated Lie Algebra registration algorithm (32). The MR images were processed using field bias correction to correct for nonuniform fields and were then segmented into gray matter (GM), white matter, and cerebrospinal fluid sections using tissue probability maps based on the International Consortium of Brain Mapping template. Individual GM images were normalized to the Montreal Neurological Institute template with a 1.5 × 1.5 × 1.5 mm^3^ voxel size and modulated for GM volumes. All normalized GM images were smoothed with a Gaussian kernel of 8 mm full width at half maximum. Consequently, the size of the input images for the proposed model was 121 × 145 × 121 voxels.

### 2.3 Cycle generative adversarial networks

The CycleGAN algorithm, a generative AI method that has been actively used in recent years for style transformation, was used in this study (33). This algorithm simultaneously learns to generate Style B from Style A, and Style A from Style B using two style datasets. In addition, whether the image is real or fake is discriminated. This enables style transformations without the need for supervised data. The GAN learns to progressively generate high-resolution images through these competitive processes. Figure 1 shows the schematic workflow of the proposed method. In this study, we constructed a model to transform images by learning two styles: MRI of HS and MRI of SZ. Using the learned model, we transformed HS into SZ and investigated how the brain regions changed pre- and post-transformation.

**Figure 1:**
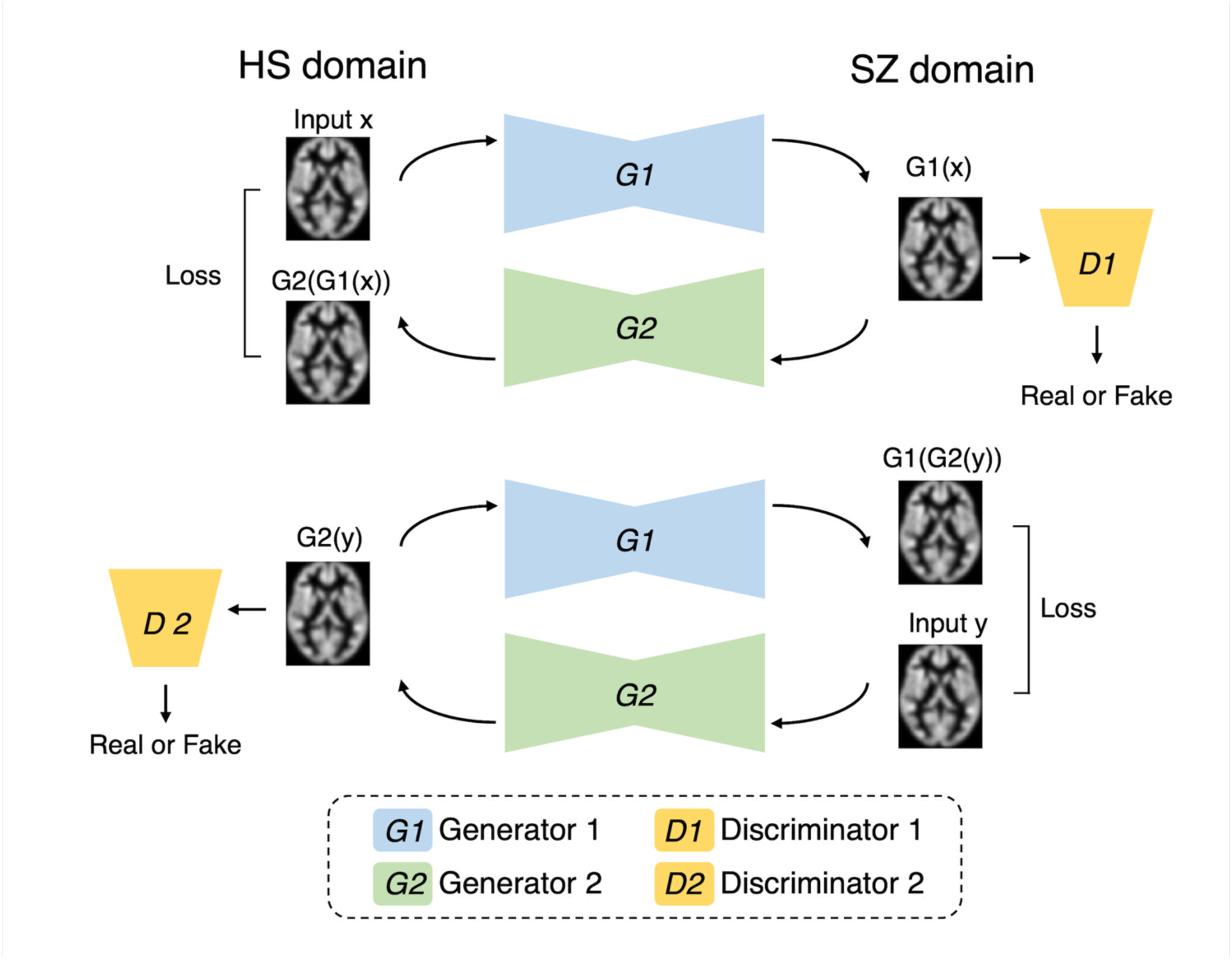
Our cycle generative adversarial network The model is designed to enable the transformation between domains of healthy subjects (HSs) and patients with schizophrenia (SZ). Generator 1 (G1) is responsible for transforming HS into SZ, whereas Generator 2 (G2) performs the conversion from SZ back to HS. Discriminator 1 (D1) discriminates between real SZ images and virtually generated SZ-like images. Similarly, Discriminator 2 (D2) discriminates between real HS images and synthetic HS-like images. The loss function is configured to optimize each component.

Figure 1 shows the proposed CycleGAN architecture. A 3D brain image was input into the proposed model to contain more spatial information, and the background area was cropped as much as possible (the voxel sizes were 96, 120, and 104). In the learning phase, the training HS MRI was input to Generator 1 (HS to SZ) to generate an SZ MRI (called virtual SZ). This virtual SZ was input to Generator 2 (SZ to HS) to generate an HS MRI (called reconstructed HS). Similarly, the training SZ MRI data were fed to the two generators in reverse order to generate a virtual HS and a reconstructed SZ. Next, two discriminators were used to judge the reality of the virtual and reconstructed images.

The network of Generators 1 and 2 consisted of U-Net (34), which is an autoencoder with skip connections. In this study, we proposed a U-Net model consisting of six consecutive 3D convolutional blocks (three encoding blocks and three decoding blocks) with instance normalization and rectified linear unit (ReLU) activation. The encoding blocks consisted of two convolutional layers and one pooling layer. The decoding block consisted of three transposed convolutional layers. Additionally, six layers of ResBlock were added to the intermediate layer of U-Net, which was used in the Residual Network to learn the residual function between the inputs and outputs of the layers (35). This structure is often used in GANs (2,36,37). The discriminator consisted of five blocks, including a 3D convolution layer with instance normalization and leaky ReLU activation. The proposed CycleGAN loss function comprises two parts: adversarial and cycle consistency losses. Adversarial loss is designed to optimize the generator’s ability to produce images that are indistinguishable from those belonging to the target domain by the discriminator. Generator *G_1_* transforms an image in domain *X* into domain *Y*, where *D_Y_* represents the discriminator for domain Y:

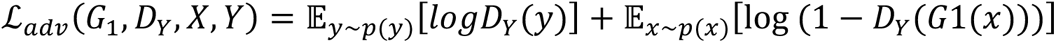

In addition, the same formulation applied to generator *G_2_* transforming an image in domain Y into domain X, with *D_X_* as the corresponding discriminator.

The cycle consistency loss ensures that the image is transformed back to its original domain and then back. This loss component is given by the following:

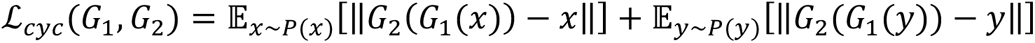

The network structure was explored preliminarily based on previous experiments (38,39). The details of the architecture of our framework are shown in Supplementary Figure 1.

We conducted the experiments in Python 3.8 using PyTorch v.1.9.1 library (40). Our network was implemented on a workstation running a 64 Gigabytes NVIDIA Quadro RTX 8000 GPU.

### 2.4 Verification of generated virtual schizophrenia brain

In this study, we used a trained CycleGAN model to generate virtual SZ MRI images of HSs. Subsequently, we analyzed the different regions of volume pre- (original HSs) and post- (virtual SZ converted from HSs) transformations and verified whether the results were consistent with the brain changes due to SZ indicated in previous case-control neuroimaging studies.

For verification, we performed voxel-based morphometry (VBM) using SPM12 (41,42). VBM is a method for comparing brain GM volumes from segmented MRI images using statistical parametric mapping to identify and infer region-specific differences. Standard and optimized VBM techniques have been used to detect psychiatric and neurological disorders (43–48). Whole-brain voxel-wise t-tests were performed, and paired t-tests were specifically employed for pre- and post-transformation comparisons. To account for potential scaling differences between pre- and post-transformation brain images, global scaling was applied to normalize the overall image intensity of each image. Correction for multiple comparisons was conducted at a combined voxel level of P < 0.001.

To ensure the reproducibility of the generation, we additionally assessed any variations in the accuracy of age prediction between pre- and post-transformations. Owing to the ultrahigh dimensionality of the brain images, principal component analysis was performed for dimensionality reduction, retaining all 71 dimensions corresponding to our sample size. Subsequently, fivefold cross-validation and linear regression were used to derive age predictions. To compare the age values predicted from the brain images pre- and post-transformation, a t-test was applied. The significance level was set at P < 0.05.

### 2.5 Brain alteration simulations using a schizophrenia brain generator

Two experiments were conducted to verify whether the developed generator can simulate brain alterations. The first involved simulating the comorbidities of the disease (Figure 2A). We applied the SZ brain generator to an independent dataset called ABIDE. This approach enables the virtual generation of an image depicting individuals with both ASD and SZ. Comparative analyses were performed between TD individuals and those with ASD as the baseline, TD individuals with SZ (TD + SZ), individuals with ASD and SZ (ASD + SZ), and individuals with ASD and ASD + SZ. The method used VBM to examine the variations in brain regions.

**Figure 2:**
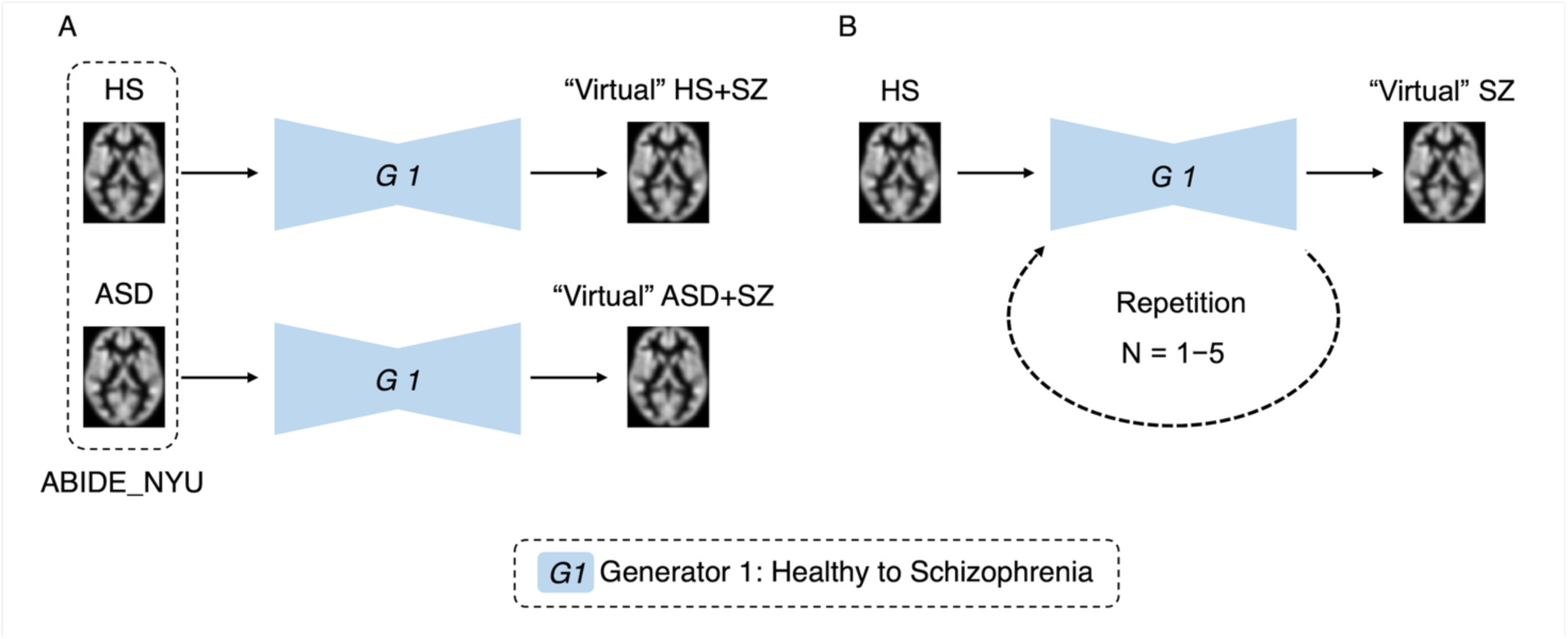
Simulation experiments by generative brain images Figure A is an experiment simulating disease comorbidity. The autism spectrum disorder and typically developing data were used to generate virtual images with a schizophrenia transducer. Figure B is an experiment simulating disease progression. Images were generated using one to five repetitions of the schizophrenia brain generator.

In the second experiment, we simulated disease progression, as shown in Figure 2B. We repeatedly applied the trained generator to the images and compared the results with those of the original images to validate the changes. Our analysis aimed to ascertain the effectiveness of this method for simulating brain alterations associated with disease progression. To confirm that the original individual characteristics were retained after repeated transformations of brain images, age predictions were made using each transformed image. An analysis of variance (ANOVA) was used to confirm the absence of significant differences in these predictions. The significance level was set at P < 0.05.

## 3 Results

### 3.1 Generation of virtual schizophrenia brain

Following an adequate learning process, our model was able to generate brain MRI images qualitatively (Supplementary Figure 2). We performed VBM analysis pre- and post-transformation to confirm that the model generated from MRI of HSs to MRI of patients with SZ captured the characteristics of schizophrenic brain structures. VBM analysis confirmed the regions of volume reduction after transformation to an SZ-like state, including the bilateral superior temporal gyrus, right middle temporal gyrus, right hippocampus, and bilateral medial frontal to anterior cingulate gyrus (Figure 3). This result is consistent with the findings of previous case-control studies on SZ (49–54), indicating that this model can reproduce the brain volume changes caused by SZ.

**Figure 3:**
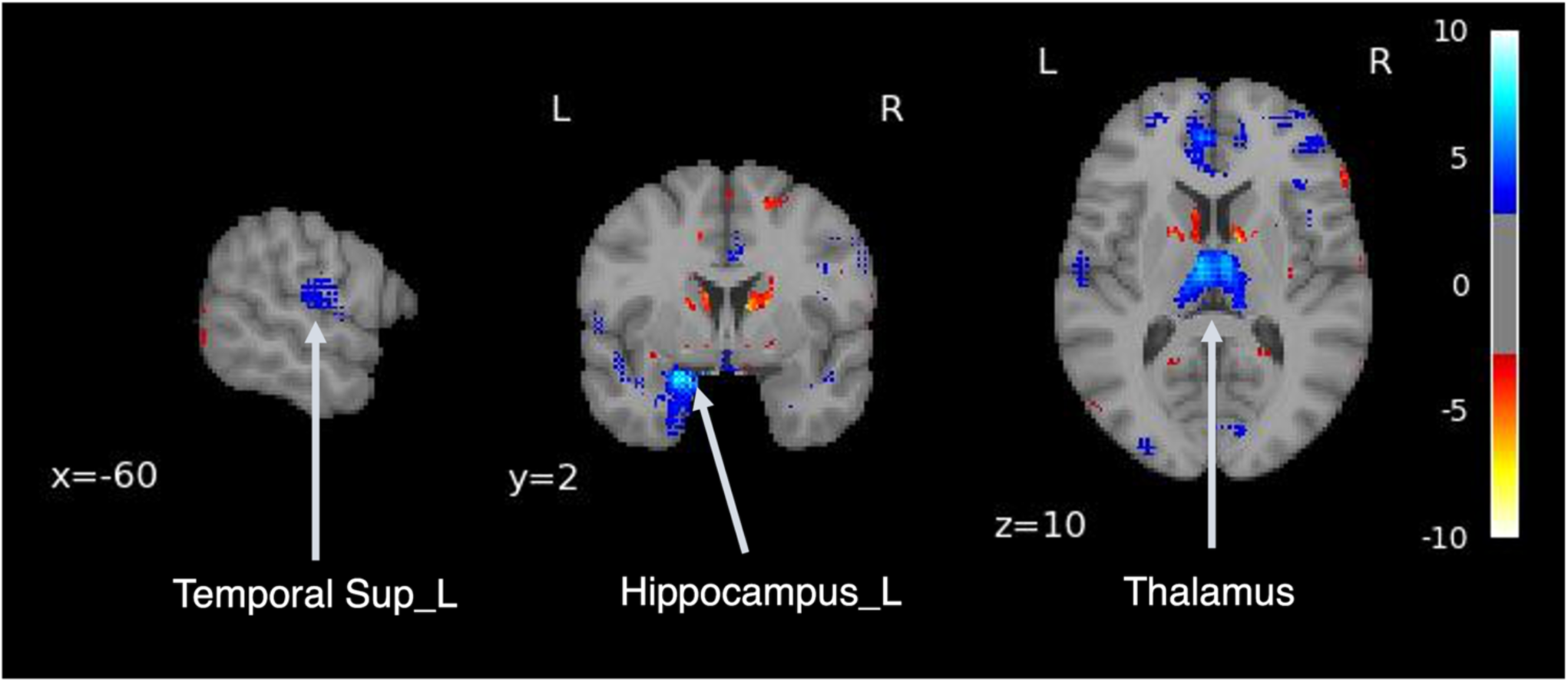
Difference in brain volume pre- and post-transformation Voxel-based morphometry analysis was performed between pre- and post-transformation magnetic resonance imaging images from healthy subjects and those with schizophrenia. Cold colors represent a decrease, and warm colors represent an increase. Temporal Sup_L, left superior temporal gyrus; SupraMarginal_L, left supramarginal gyrus.

In addition, age prediction was performed using brain images obtained pre- and post-transformation, and there was no significant difference in the predicted values (P = 0.598). In addition to the t-test, the effect size was calculated using Cohen’s d, which revealed a small effect (d = 0.089) (Supplementary Figure 3).

Using a generator model that transforms the MRI of HSs into the MRI characteristics of patients with SZ, this model was applied to MRI images, and the volume differences of brain regions pre- and post-transformation were examined. After applying SZ brain generator images, volume reduction was observed in the bilateral superior temporal gyrus, right middle temporal gyrus, right hippocampus, and bilateral medial frontal gyrus to the anterior cingulate gyrus.

### 3.2 Simulation analysis of virtual schizophrenia with autism spectrum disorder brain

We performed a simulation using a trained SZ MRI generator. We generated virtual MRI images of ASD with SZ (ASD + SZ) by transforming them from MRI images. In Figure 4A, the cold-colored areas represent regions where the volume was reduced when ASD was comorbid with SZ. We confirmed volume reduction, mainly in the bilateral temporal lobes and insular cortex.

**Figure 4:**
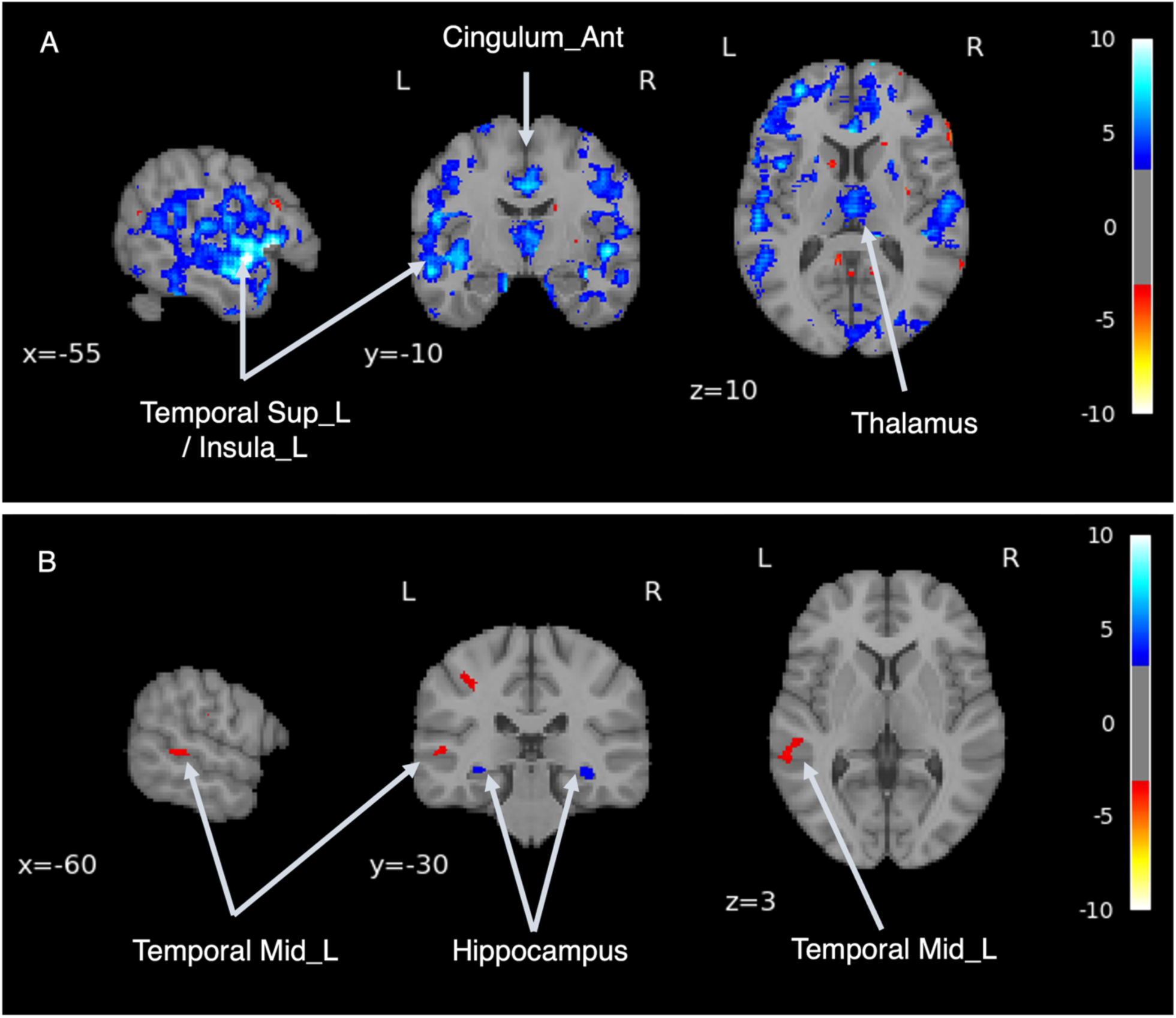
Disease comorbidity simulation Figure A shows the volume differences before and after the application of the schizophrenia brain generator, which enabled the generation of an image of autism spectrum disorder (ASD) combined with schizophrenia (SZ). Figure B shows the volume differences between the virtual SZ-like images with and without ASD.

**Figure 5:**
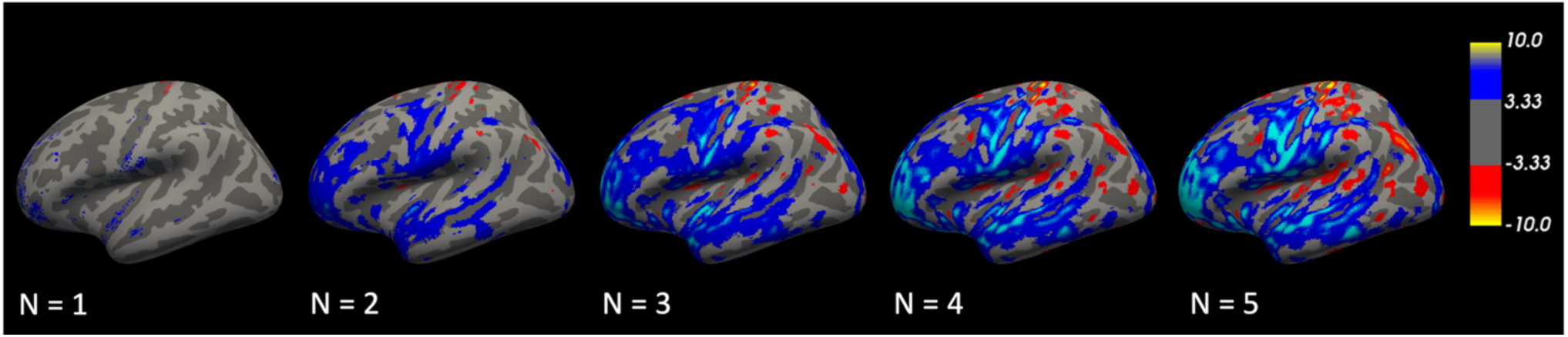
Repetitive transformations The volume differences were relatively localized at first, but gradually became more extensive, especially in the temporal lobe, by the fifth transformation.

Next, we performed a simulation to examine whether there were differences in brain structure between individuals diagnosed with SZ with and without background ASD. Virtual SZ MRI images were generated from ASD and TD MRI images using an SZ brain generator. Depicted in Figure 4B are regions where the cold regions show a volume reduction in the virtual SZ generated from the ASD (ASD + SZ) compared with the virtual SZ generated from the TD (TD + SZ). Volume reduction was observed bilaterally in the hippocampus. The warm regions showed volume increases in the ASD + SZ images. An increase in volume was observed in the left middle temporal gyrus.

### 3.3 Simulation analysis of repetitive transformations

We hypothesized that repeated transformations of brain images could potentially delineate the evolutionary trajectory of an ailment. Employing the VBM, an investigation was undertaken on both the original images and the nth iterated rendition (n = 1–5). The results showed that the region of difference expanded with each repeated transformation. Although it was initially relatively localized, it progressively became widespread and centered on the temporal lobe during the fifth transformation.

To confirm that the original individual characteristics were retained after repeated transformations of the brain images, age predictions were conducted using each transformed image.

Fivefold cross-validation and linear regression were used to predict age from repeatedly transformed brain images. The differences in prediction accuracy were evaluated using ANOVA. The results showed no significant differences in age prediction (P = 0.932, η²p = 0.01) (Supplementary Figure 4). This suggests that the original individual characteristics are preserved after multiple transformations.

## 4 Discussion

In this study, we introduced generative AI capable of depicting structural differences in the brain resulting from psychiatric disorders. Specifically, we developed a generative AI to transform brain images into SZ images. Our results confirm the potential of the model for facilitating several simulation experiments related to psychiatric disorders.

Our model was qualitatively successful in transforming the MRI images of HSs into images resembling those of patients with SZ. Subsequent VBM analysis confirmed the alignment of the model with previously established SZ studies. Patients with SZ exhibit structural anomalies in the superior temporal gyrus, thalamus, and hippocampus compared with healthy individuals, a finding that corroborates our results (53–55). This study offers a compelling solution to the problem of insufficient neuroimaging data by effectively generating data that capture the characteristics of SZ-related brain changes (56).

Subsequent simulations using a trained SZ brain generator extended the investigation to individuals with both SZ and ASD. SZ and ASD are distinct disorders with unique clinical profiles and natural history. However, ASD carries a significantly higher risk, three–six times, of developing SZ than TD individuals (57,58). Recent studies have indicated a convergence between SZ and ASD. To investigate this intricate relationship, we conducted a virtual brain simulation and proposed a new hypothesis. This exploration revealed volume reduction patterns, primarily concentrated in the bilateral temporal lobes and insular cortex, which are characteristic of comorbidities. This observation suggests that the model captures distinct structural changes specific to this subgroup, thereby demonstrating its potential to unravel the complex interplay between different psychiatric conditions.

Further simulations were performed assuming a retrospective study. In this study, we generated brain images of patients with SZ with and without ASD and examined whether it was possible to analyze the differences in their structures. The distinct volume reductions observed in the bilateral hippocampus in the ASD + SZ group indicate a potential structural divergence associated with the comorbidity. Conversely, the volume increase in the left middle temporal gyrus in the same group offers an interesting avenue for understanding unique structural variations in this population. Zheng et al. reported that the higher the autistic traits, the lesser the improvement in psychiatric symptoms and life functioning after a year (59). Therefore, it is important to determine the presence or absence of ASD in the context of SZ to predict the prognosis and determine the course of treatment. The proposed model can provide decision support for treatment strategies.

The repeated transformation approach, which was designed to explore the evolutionary trajectory of brain changes, provides novel insights into ailment progression. The expansion of the difference region with each repeated transformation, culminating in a widespread pattern centered on the temporal lobe, underscores the model’s ability to capture and magnify the cumulative effects of structural alterations. A meta-analysis of longitudinal studies on SZ revealed that patients with SZ exhibited significantly higher volume loss over time (49). This loss included the entire cortical GM, left superior temporal gyrus, left anterior temporal gyrus, and left Heschl’s gyrus. These findings are consistent with the simulation results generated using the proposed model. This repeated approach can potentially aid in elucidating the progressive nature of the impact of the disorder on the brain structure.

Investigation of the preservation of the original individual characteristics after repeated transformations brought an essential dimension to the study. By evaluating age predictions across the transformed images, this study established the robustness of the model in retaining individual-specific features. The absence of statistically significant differences in age predictions reinforces the credibility of repeated transformations in preserving the key characteristics of the original images.

Despite these promising findings, it is crucial to acknowledge the limitations of this study. One limitation is the reliance on a simulated data approach, which may not fully capture the complexity of real-world brain structural variations. Additionally, even if there is no difference in the accuracy of age prediction using the generated brain images, it may not cover the entire range of demographic factors that could influence brain structure. Further validation with larger and more diverse datasets and comparisons with other methodologies can enhance the generalizability of the results.

In conclusion, this study demonstrated the potential of the developed model to capture and simulate brain structural alterations associated with SZ and its comorbidity with ASD. These findings provide a foundation for exploring the mechanisms underlying these conditions and their interconnections.

## 6 Author Contributions

HY and YY conceived and designed the study. HY conducted the deep learning experiments and analyzed the data. HY and YY drafted the manuscript. GS and MS provided the critical revisions. All the authors contributed to and approved the final version of the manuscript.

## Data Availability

All data produced in the present work are contained in the manuscript.

https://fcon_1000.projects.nitrc.org/indi/retro/cobre.html

https://fcon_1000.projects.nitrc.org/indi/abide/

## 8 Acknowledgments

We thank all the authors of the included studies. We also would like to thank Editage (www.editage.com) for English language editing.

## 10 Data Availability Statement

All data generated or analyzed in this study are included in the published article. Primary data were obtained from public databases, including the Center for Biomedical Research Excellence (COBRE; http://fcon_1000.projects.nitrc.org/indi/retro/cobre.html) and Autism Brain Imaging Data Exchange (ABIDE; http://fcon_1000.projects.nitrc.org/indi/abide/).

---

**Supplementary Table 1:** Demographic and clinical information.

| Characteristic | COBRE (n = 142) |  | ABIDE_NYU (n = 184) |  |
| --- | --- | --- | --- | --- |
|  | SZ | HS | ASD | HS (TD) |
|  | n=71 | n=71 | n=79 | n=105 |
| Age, mean (SD), year | 39.9 (8.2) | 36.5 (8.5) | 14.5 (7.0) | 15.8 (6.3) |
| Sex, Male/Female | 58/13 | 50/21 | 68/11 | 79/26 |
| FIQ, mean (SD) | 97.8 (16.8) | 106.8 (11.2) | 107.9 (16.6) | 113.2 (13.1) |
| VIQ, mean (SD) | 103.2 (16.3) | 114.7 (11.6) | 105.8 (16.1) | 113.1 (12.6) |
| PIQ, mean (SD) | 99.8 (16.9) | 112.1 (11.6) | 108.8 (17.4) | 110.1 (13.7) |
| CPZE, mean (SD) | 407.6 (1072.6) |  |  |  |
| PANSS, mean (SD) | 58.8 (13.8) |  |  |  |
| ADOS, mean (SD) |  |  | 11.30 (4.1) |  |

**Supplementary Figure 1:**
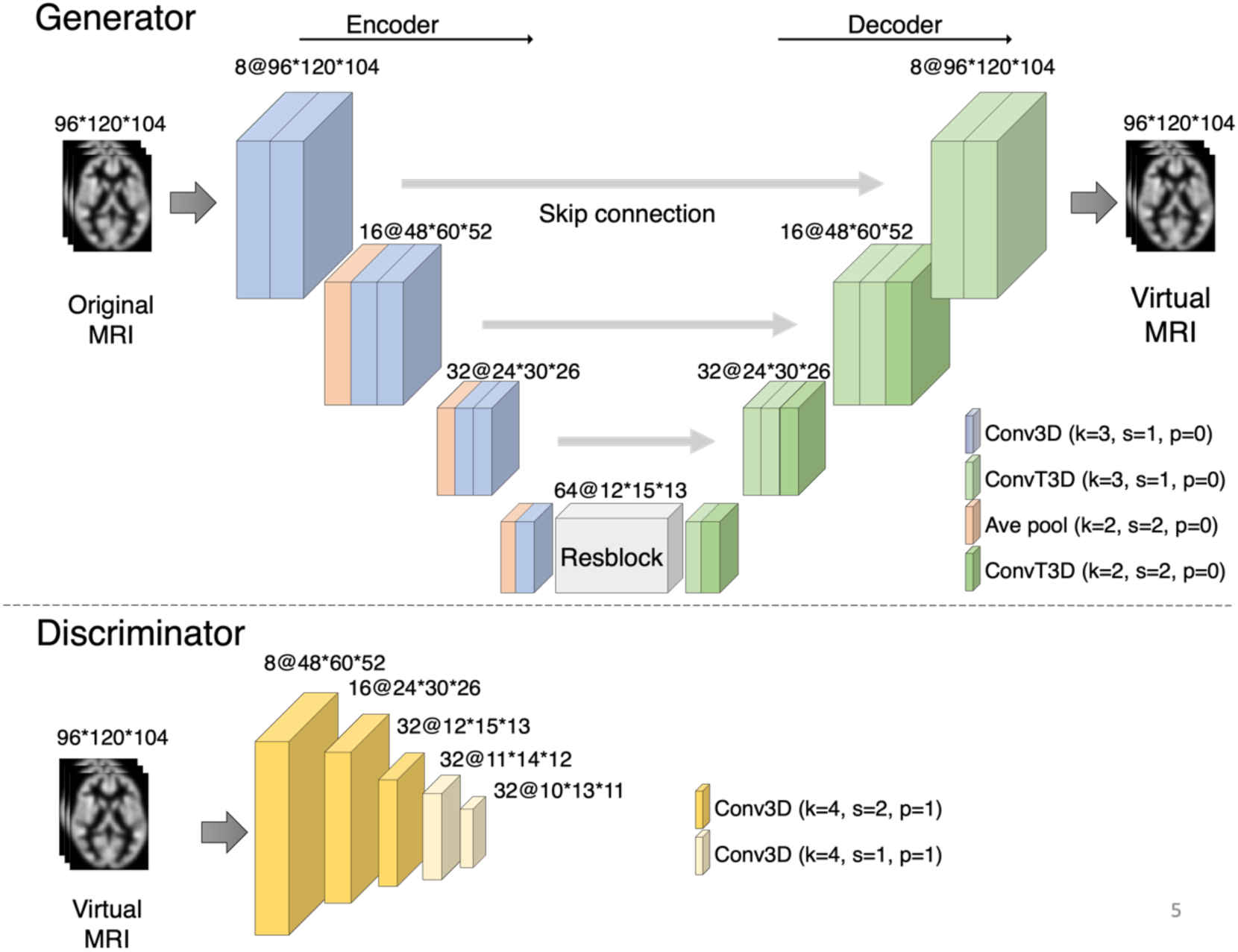
Our proposed CycleGAN architecture. The numbers describe the number of channels and the size of the images. The generator consists of an encoder and a decoder, and the bottleneck contains a ResBlock. The encoder and decoder were designed as a U-Net with skip connections. The discriminator, a convolutional neural network, is trained to discriminate between the generated images and the ground truth.

**Supplementary Figure 2:**
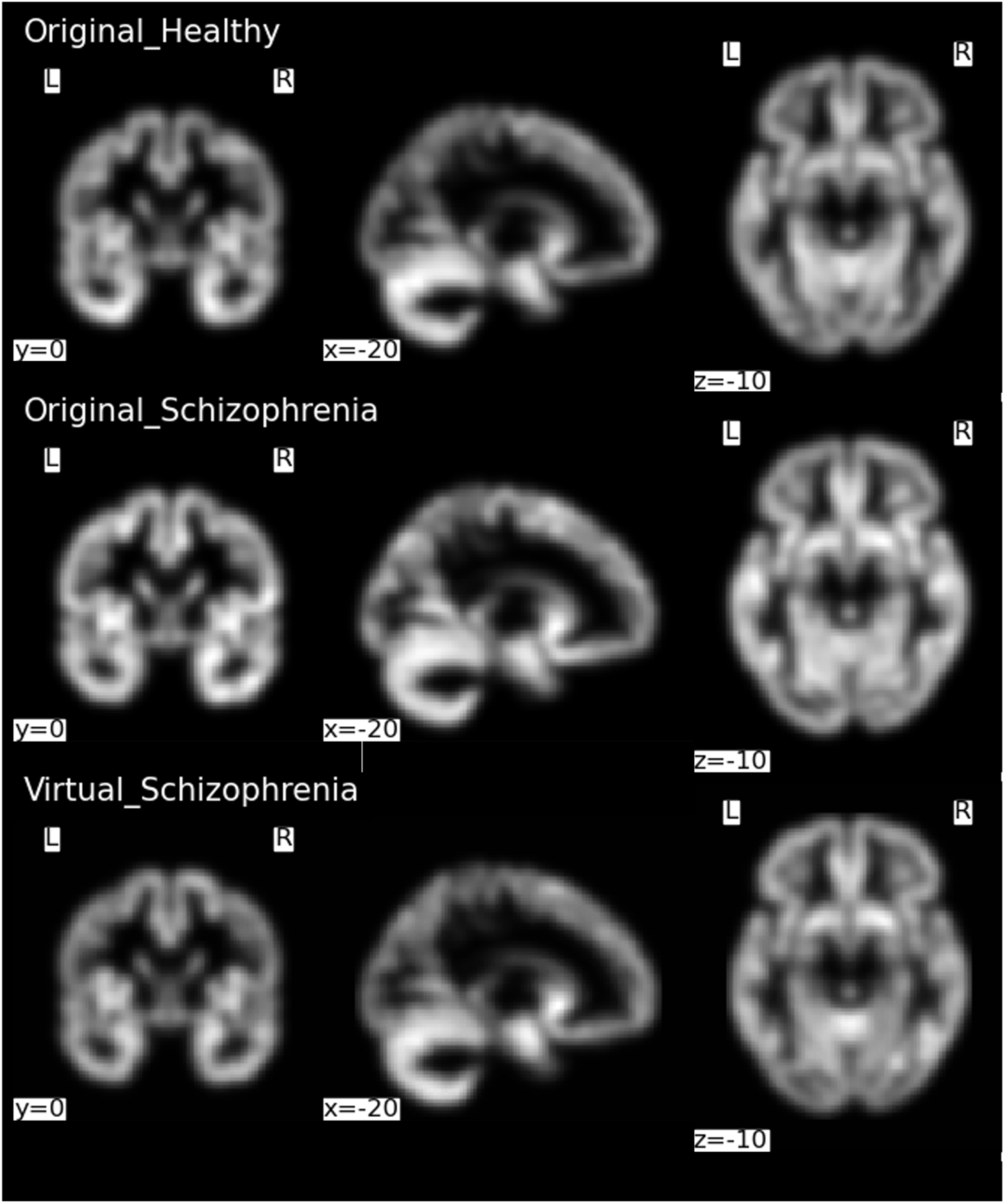
Comparison of MRI images of healthy subjects, schizophrenia patients, and generated virtual schizophrenia MRI images. MRI images of a healthy subject (top row), a real schizophrenia patient (middle row), and a virtual schizophrenia patient (bottom row). Coronal (left), sagittal (middle), and axial (right) views are shown, respectively. A virtual schizophrenia was generated from a healthy subject. It appears qualitatively no different from the real image.

**Supplementary Figure 3:**
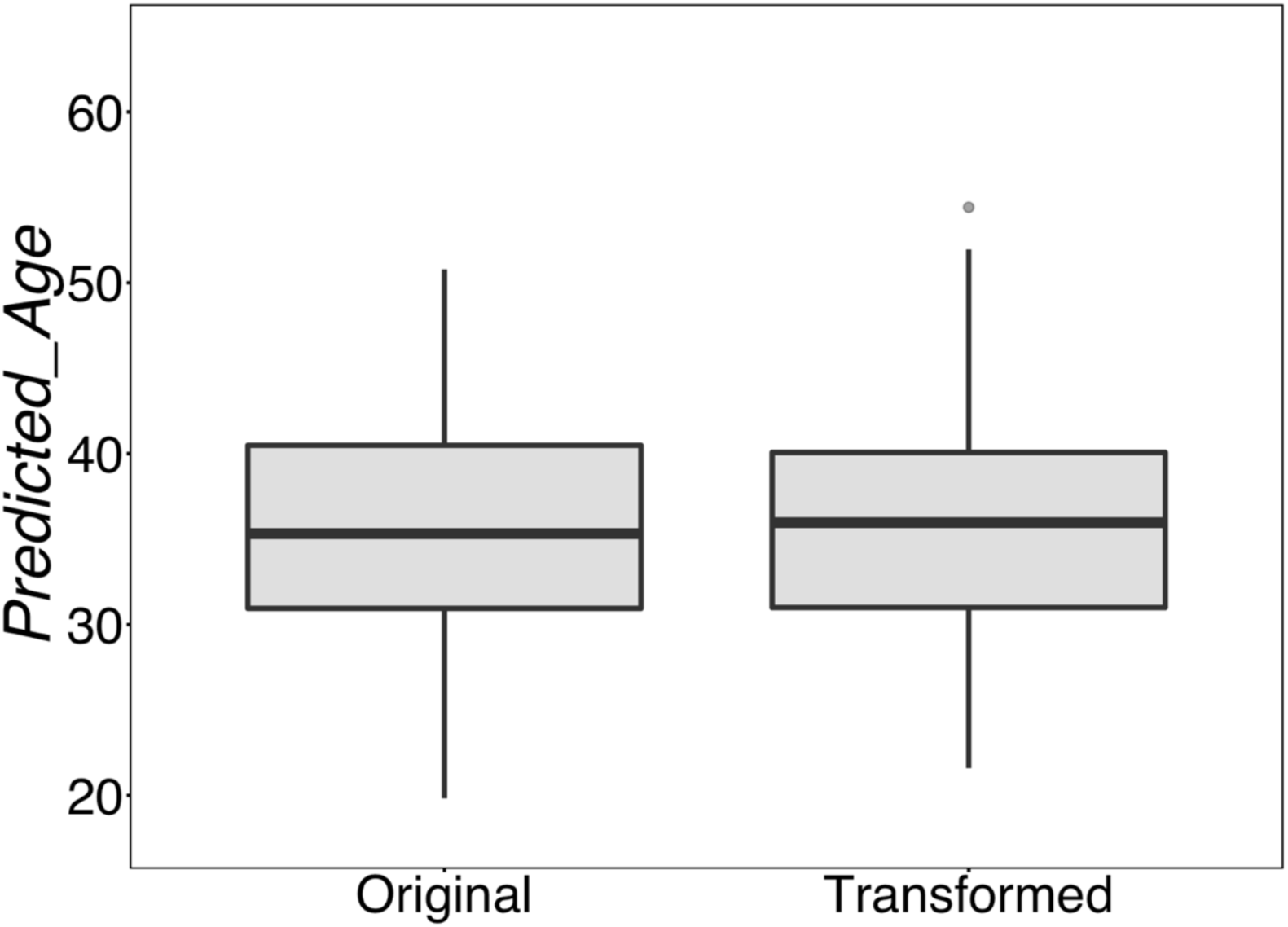
Comparison of age predictions pre- and post-transformation Healthy subject brain MRI was transformed to schizophrenia brain MRI and linear regression was performed for age predictions pre- and post-transformation, respectively. There were no significant differences in predictive values pre- and post-transformation.

**Supplementary Figure 4:**
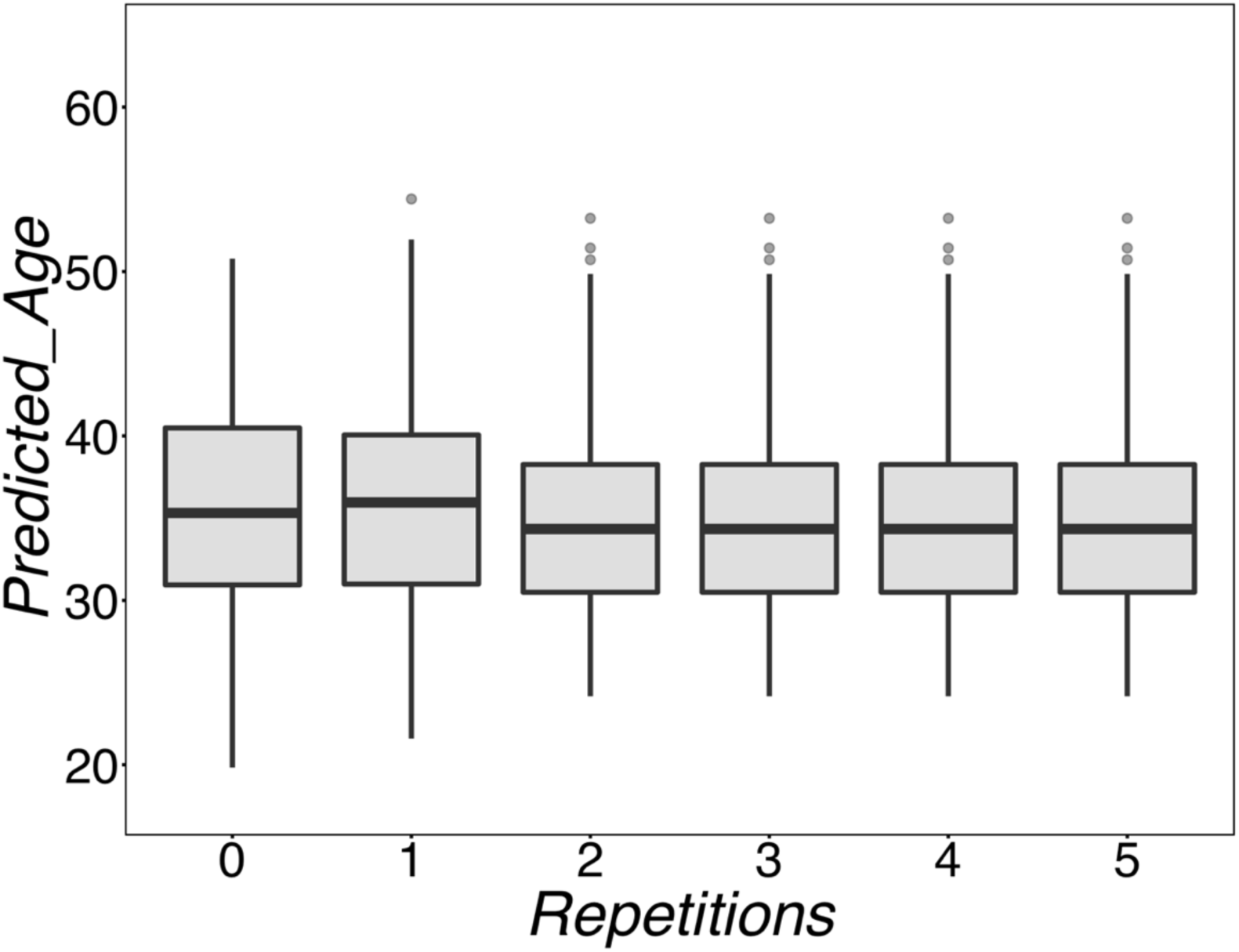
Comparison of age prediction by repeatedly transformed brain images Repeated transformation experiments were performed using the schizophrenia transformer. A linear regression was performed to predict age by each transformed image, and an ANOVA showed no significant difference in predicted values.

